# Oxidative stress induced liver damage in dengue is exacerbated in those with obesity

**DOI:** 10.1101/2025.03.18.25324170

**Authors:** Heshan Kuruppu, Maneshka Karunananda, Chandima Jeewandara, Laksiri Gomes, D.M.C.B Dissanayake, Chathura Ranatunga, Padukkage Harshani Chathurangika, Nushara Senatilleke, Navanjana Warnakulasuriya, Rivindu H. Wickramanayake, Ananda Wijewickrama, Damayanthi Idampitiya, Graham S Ogg, Gathsaurie Neelika Malavige

**Affiliations:** Allergy, Immunology and Cell Biology Unit, Department of Immunology and Molecular Medicine, University of Sri Jayewardenepura, Nugegoda, Sri Lanka; National Institute of Infectious Diseases, Angoda, Sri Lanka; MRC Translational Immune Discovery Unit, MRC Weatherall Institute of Molecular Medicine, University of Oxford, United Kingdom, University of Oxford, Oxford, United Kingdom

## Abstract

**Background:** Obesity and diabetes are risk factors for severe dengue. As there are limited data on the association of obesity with liver dysfunction and oxidative stress in patients with acute dengue, we investigated liver dysfunction associated with obesity, oxidative stress and inflammatory markers, in a large cohort of patients with varying severity of acute dengue.

**Methods:** 577 adults dengue patients with acute disease, presenting with a duration of illness ≤ 4 days, were enrolled and followed up from admission to discharge, with clinical and laboratory features recorded. Aspartate transaminase (AST), alanine transaminase (ALT), C-reactive protein, ferritin, 4-hydroxynonenal (4-HNE) and malondialdehyde (MDA) levels were measured, along with the height, weight and waist circumference.

**Results:** AST, ALT, CRP and ferritin levels were significantly higher in patients with central obesity (waist circumference of ≥80cm in women or ≥90cm in men) compared to leaner individuals. ALT and CRP levels were also significantly higher in patients with a BMI of ≥ 23.9 kg/m^2^. 4-HNE levels significantly increased with the rise in AST levels and with ALT levels although not significant. In contrast, MDA levels gradually decreased with the rise in AST levels and ALT levels. There were no differences in 4-HNE and MDA levels in relation to clinical disease severity. However, MDA levels were significantly higher in younger individuals, and leaner individuals with a normal BMI. Furthermore, MDA levels inversely correlated with serum ferritin levels, while AST, ALT and CRP levels significantly correlated ferritin levels.

**Conclusions:** 4-HNE and MDA which are markers of lipid peroxidation, appear to play different roles in the pathogenesis of dengue, which should be further investigated for identification of therapeutic targets for treatment of dengue.

## Background

Dengue has been named a pathogen with a pandemic potential in World Health Organization R&D blueprint, due to its rapid spread causing outbreaks in 176 countries [1, 2]. Although most dengue viral (DENV) infections result in asymptomatic illness, some individuals develop symptomatic illness which may manifest as a mild illness in the form of an undifferentiated febrile illness or dengue fever (DF). However, some individuals develop severe disease manifestations such as plasma leakage, bleeding and organ dysfunction [3]. It is estimated that 36.8% of hospitalized patients develop plasma leakage in the form of pleural effusion and ascites, which left untreated can lead to shock resulting in fatalities [4]. Another important complication in patients with dengue is liver dysfunction. 60 to 90% of patients with dengue are reported to have liver involvement [5]. Further, the liver transaminases, aspartate transaminase (AST) and alanine transaminase (ALT) levels during the febrile phase have shown to be useful predictors of the occurrence of severe dengue [6].

Acute liver failure (ALF) in dengue is defined by either AST or ALT levels >1000 IU/L [3]. ALF has been reported in 0.4% to 36.4% of patients with dengue, while it has been reported in 53.6% to 71% of patients who have succumbed to their illness [5]. Systematic reviews confirm ALF having poor prognosis in patients with dengue, reporting 47% mortality rates in such patients [7]. While elevated AST and ALT levels are seen in >75% of patients with DHF [6, 8], the extent of the rise of liver transaminases were shown to be lower in children compared to adults [5]. Further, while plasma leakage in seen in most patients with ALF, there have been reports of patients developing ALF in the absence of detectable plasma leakage [9–11]. Therefore, the pathogenesis of liver involvement during acute dengue appears to be multifactorial. Multiple factors such as hypoxic injury due to poor organ perfusion, direct hepatocyte death due to the virus and immune mediated mechanisms are thought contribute to liver dysfunction in dengue [12]. Varying degrees of hepatic necrosis, sinusoidal congestion, macrovascular and microvascular steatosis and Kupffer cell hyperplasia are seen postmortem studies in patients who succumbed to their illness [12].

Although the pathogenesis of liver dysfunction in dengue has not been extensively studied, the macrovascular and microvascular steatosis seen in fatal dengue due to liver damage, is similar to changes seen in patients with metabolic dysfunction-associated steatotic liver disease (MASLD), and non-alcoholic fatty liver disease (NAFLD). Indeed, patients with NAFLD who were infected with the DENV were significantly more likely to have higher haemoconcentration, lower platelet counts, and a prolonged hospital stay compared to those without NAFLD [13]. Oxidative stress leading to excessive production of reactive oxygen species causing mitochondrial dysfunction is thought to play a central role in the pathogenesis of MASLD [14, 15]. Recently it was shown that arginase-1, α-glutathione S transferase, and 4-hydroxyphenylpyruvate dioxygenase levels during early illness associated with the extent of liver damage [16]. These enzymes and mediators are involved in modulating cellular oxidative stress [17, 18]. Infection with the DENV induces oxidative stress, which leads to inhibition of viral replication [19, 20]. However, excessive and unregulated oxidative stress may lead to extensive liver damage in patients, who already have existing liver damage possibly in the form of MASLD/NAFLD.

The risk of severe dengue is several fold higher in those with comorbidities such as obesity and diabetes [12]. We and others recently showed that obesity is also associated with an increased risk of symptomatic illness, and hospitalization [21, 22]. Therefore, obesity associated with metabolic disease may increase the risk of liver dysfunction, possibly due to excessive and unregulated oxidative stress. To answer this question, in this study we investigated liver dysfunction in a large cohort of patients with varying severity of acute dengue and sought to explore associations of obesity, liver dysfunction, oxidative stress and inflammatory markers.

## Methods

### Recruitment of patients and classification of disease severity

A total of 577, adult patients presenting with a suspected acute dengue infection were recruited from the National Institute of Infectious Diseases, Sri Lanka, between December 2022 and December 2024, following informed written consent. They were considered to be having an acute dengue infection if they had a positive NS1 antigen test (SD Biosensor, South Korea) or by a positive DENV specific real-time PCR. All those with known chronic kidney disease or chronic liver disease were excluded from the study.

Patients were recruited during the febrile phase (≤ 4 days since onset of illness), with the first day of onset of fever being considered as the first day of illness. Liver transaminases and C-reactive protein (CRP) were measured in all patients on the day of admission (on the day of recruitment to the study). Clinical symptoms and laboratory parameters were recorded multiple times per day while in hospital. Fluid leakage was assessed daily using bed-side ultrasound scans to detect pleural effusions and ascites. Disease severity was classified according to the 2011 World Health Organization (WHO) dengue classification criteria. Accordingly, patients with a rise in the haematocrit of ≥ 20% from baseline or those with ultrasound evidence of plasma leakage were classified as having dengue hemorrhagic fever (DHF). Patients who developed shock, defined by a pulse pressure narrowing to 20 mmHg, were categorized as having dengue shock syndrome (DSS).

Based on these criteria, 163 patients were classified as having DHF, while 414 patients were categorized as having dengue fever (DF). From this cohort, a subset of 94 DF patients and 66 DHF patients were randomly selected for the assessment of oxidative stress markers, including 4-hydroxynonenal (4HNE), malondialdehyde (MDA), and ferritin levels, to represent mild (DF) and severe dengue cases (DHF), respectively.

#### Anthropometric measurements

The height and weight of the patients were measured at the time of recruitment. The weight was measured using a digital scale, while wearing light clothing and barefooted. The height was measured barefooted, using a stadiometer. Waist circumference was measured using a non-stretchable tape between the lowest rib and the iliac crest after exhalation. Those who had a body mass index of ≥ 23.9 kg/m^2^ were considered as overweight [23]. Women with a waist circumference of ≥80cm or men with a waist circumference of ≥90cm were classified as having central obesity, as previously defined [24].

#### Ethics Statement

This study was approved by the Ethics Review Committee of the Faculty of Medical Sciences, University of Sri Jayewardenepura, Sri Lanka (Ethics Application Number: 58/19). Informed written consent was obtained from all participants before enrollment in the study.

#### Quantification of 4-hydroxynonenal (4HNE) and malondialdehyde (MDA) levels

Serum samples stored at -80°C were used for the measurement of 4HNE and MDA levels using commercially available quantitative enzyme-linked immunosorbent assay (ELISA) kits (Antibodies.com, UK). Assays were performed according to the manufacturer’s instructions, with serum samples diluted at predefined ratios as per the protocol. A four-parameter logistic regression model was used to interpret the assay results. 4HNE and MDA levels were measured in serum samples obtained during the febrile phase of illness.

#### Quantification of Ferritin Levels

Ferritin levels were measured in serum samples stored at -80°C until analysis. The quantification was performed using the AutoLumo A6200 analyzer (AutoBio, China), a chemiluminescence immunoassay-based automated system. The assay was conducted according to the manufacturer’s protocol, ensuring high specificity and sensitivity for ferritin detection. Calibration and quality control procedures were performed prior to analysis to maintain assay accuracy.

#### Statistical analysis

Statistical analyses were conducted using GraphPad Prism version 10.4.1 (Dotmatics, California, USA). Since the data did not follow a normal distribution, differences in oxidative stress markers between different clinical severity groups were analyzed using the Mann-Whitney U test (two-tailed). Correlations between variables, including associations between oxidative stress markers and laboratory parameters, were assessed using Spearman’s rank-order correlation coefficient (two-tailed).

## Results

### Liver transaminases, CRP and ferritin levels in relation to clinical disease severity and obesity

As reported in many studies, both aspartate transaminase (AST) levels and alanine transaminase (ALT) levels were significantly higher in patients with DHF compared to those with DF [6, 9] (Fig 1A). The waist circumference was available in 422 patients and the BMI was available in 539 patients. Of these 422 patients, 191/422 (45.3%) were considered have central obesity and 245/539 (45.4%) were found to have a BMI ≥ 23.9 kg/m^2^. Interestingly, those who were obese or were overweight were significantly more likely to have higher AST and ALT levels (Fig 1B). Again, as seen in previous studies the CRP levels were significantly higher in those with DHF compared to DF [25, 26]. However, again as seen with the liver transaminases, the CRP levels were significantly higher in those with central obesity and in those with a BMI of ≥ 23.9 kg/m^2^ (Fig 1C).

**Figure 1:**
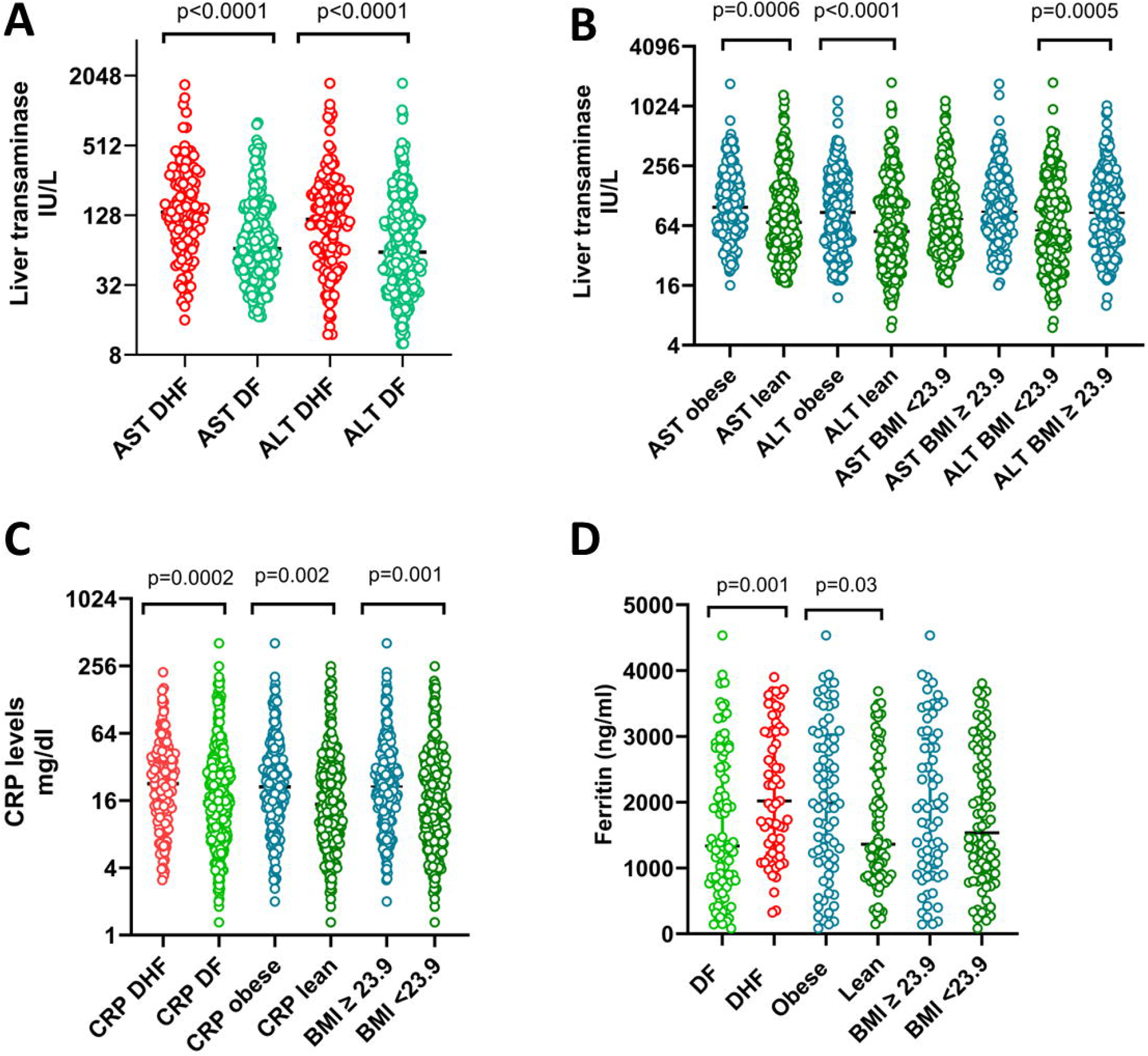
Liver transaminases, CRP and ferritin levels in relation to clinical disease severity and obesity in patients with acute dengue. Aspartate transaminase (AST) levels and alanine transaminase (ALT) levels were measured in patients with DHF (n=163, red symbols) and DF (n=414, green symbols). AST and ALT levels were also analyzed in those who had central obesity (n=191, blue) and lean individuals (n=231, green) and in those with a ≥ 23.9 kg/m2 (n=245, blue) and those with a BMI of <23.9 (n=294, green) (B). CRP levels were also measured in those with DF and DHF, those who had central obesity and lean individuals and those with a ≥ 23.9 kg/m2 and those with a BMI of <23.9 (C). Ferritin levels were measured in those with DHF (n=63, red symbols) and DF (n=88, green symbols), in those with central obesity and lean individuals and in those with a ≥ 23.9 kg/m^2^ and those with a BMI of <23.9 kg/m^2^ (D). The Mann-Whitney U test (two tailed) was used to calculate the differences between different cohorts. The error bars indicate the median and the interquartile ranges.

Ferritin has shown to be elevated in patients with severe dengue with 63% of patients reporting levels > 2000ng/ml in some studies [27, 28]. We measured ferritin levels in 88 patients with DF and 63 patients with DHF. We too observed that ferritin levels were indeed significantly higher in patients with DHF compared to DF (Fig 1D). We found that 29 (32.9%) patients with DF and 32 (50.8%) of patients with DHF had ferritin levels > 2000ng/ml. Again, as observed with liver enzymes and CRP levels, ferritin levels were significantly higher in obese individuals compared to lean individuals, although no difference was found in those with high and low BMIs (Fig 1D).

#### Association of markers of oxidative stress with liver transaminase levels

4-hydroxy-2-nonenal (4-HNE) has been identified as a marker and a causative factor of oxidative stress [29]. While malondialdehyde (MDA) is also a marker of oxidative stress, it is considered to be less toxic, yet more mutagenic than 4HNE [30]. Therefore, we measured the levels of 4HNE and MDA in 94 patients with DF and 66 patients with DHF to understand their potential association with the extent of liver damage. We found that 4-HNE levels gradually and significantly increased with the rise in AST levels (Fig 2A). Although 4-HNE levels also increased with the rise in ALT levels these differences were not significant. In contrast, we observed that MDA levels gradually decreased with the rise in AST levels and ALT levels (Fig 2B). Those with ALT levels between 41-400 had significantly lower MDA levels than those with normal ALT levels.

**Figure 2:**
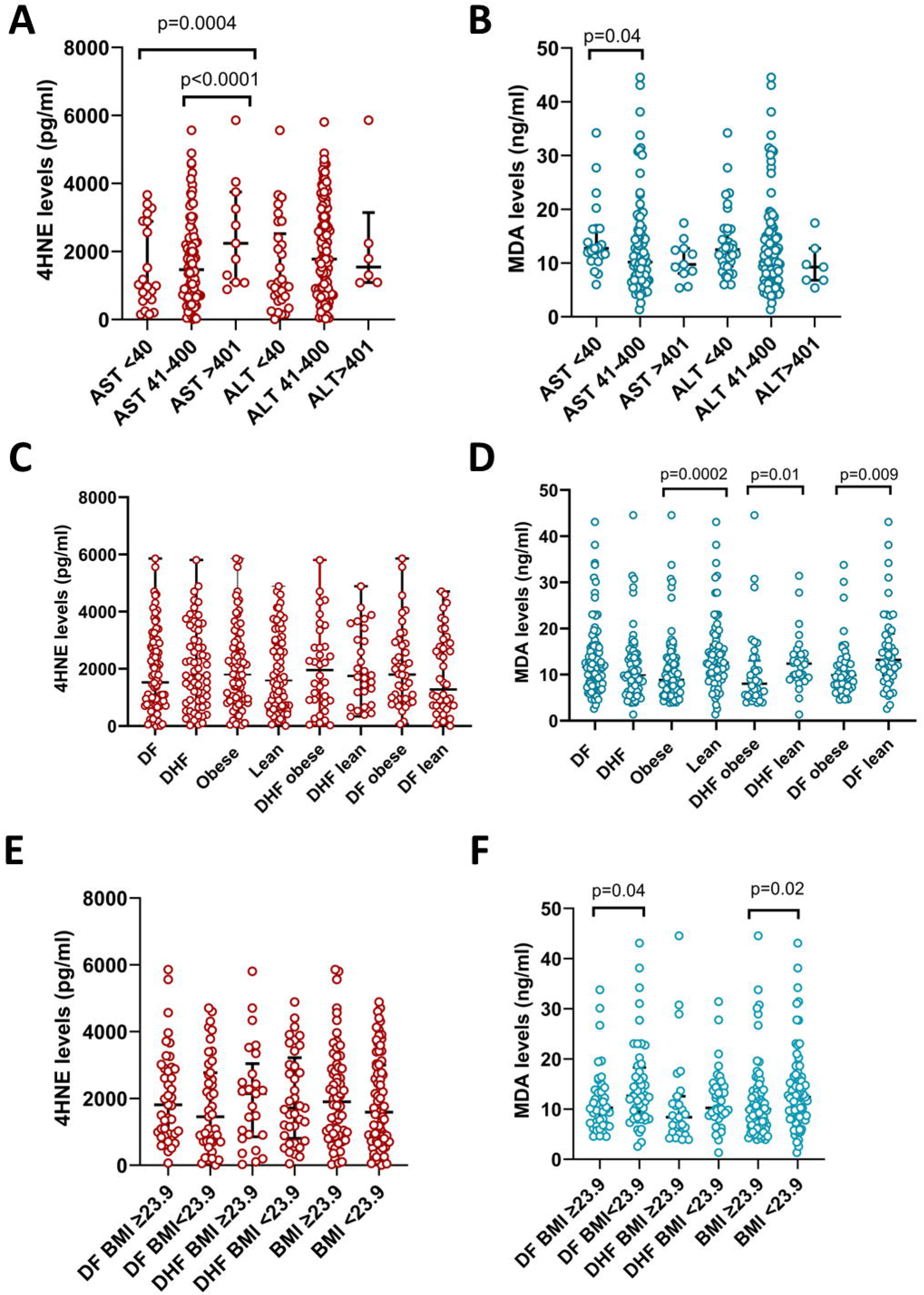
Association of markers of oxidative stress with liver transaminase levels. 4HNE (A, dark red) and MDA levels (B, teal) were compared in patients with AST levels <40, 41-400 and >400 IU/L. 4HNE (C) and MDA (D) was also compared in patients with DF (n=94) and DHF (n=66) those with central obesity and lean individuals and in those with a ≥ 23.9 kg/m^2^ (E) and those with a BMI of <23.9 kg/m^2^ (F). The Mann-Whitney U test (two tailed) was used to calculate the differences between different cohorts. The error bars indicate the median and the interquartile ranges.

As we observed that obese individuals and overweight individuals had higher liver transaminases and disease severity, we sought to investigate the changes in 4-HNE and MDA in relation to clinical disease severity and obesity. There was no difference in 4-HNE levels in those with DF compared to DHF and no difference in relation to central obesity (Fig 2C). Similarly, although there was no significant difference in MDA levels in those with DF and DHF, MDA levels were significantly higher in lean individuals, and also in lean patients with DF and DHF (Fig 2D). Again, there was no difference in 4-HNE levels with BMI > or < 23.9 kg/m^2^ (Fig 2E). However, MDA levels were significantly higher in those with a BMI of <23.9 kg/m^2^ compared to those with a BMI of ≥ 23.9 kg/m^2^ (Fig 2F). MDA levels inversely correlated with the waist circumference of individuals (Spearmans R=-0.34, p<0.0001) and with the BMI (Spearmans R=- 0.22, p=0.004), whereas 4-HNE did not correlate with any of these parameters, nor with MDA levels.

#### Relationship of markers of oxidative stress with age

Previous studies carried out in a smaller number of patients, where the study population was children or younger individuals showed that both 4HNE and MDA levels were higher in patients with acute dengue and correlated with disease severity, while some studies showed no significant differences [31–33]. As we observed that 4-HNE was associated with a rise in liver transaminases, we found that MDA levels inversely correlated with the liver transaminases and were significantly higher in lean individuals compared to obese individuals. Therefore, we proceeded to understand any age specific differences between 4-HNE and MDA levels in our cohort of patients. We found that there was no difference between 4-HNE levels in different age groups (Fig 3A), while MDA levels were significantly lower in those aged ≤ 25 years compared to those aged 41-55 years or those aged >56 years (Fig 3B). MDA levels inversely correlated with the age (Spearmans’ R= -0.27, p=0.0006), whereas, no such correlation was seen with 4- HNE levels.

**Figure 3:**
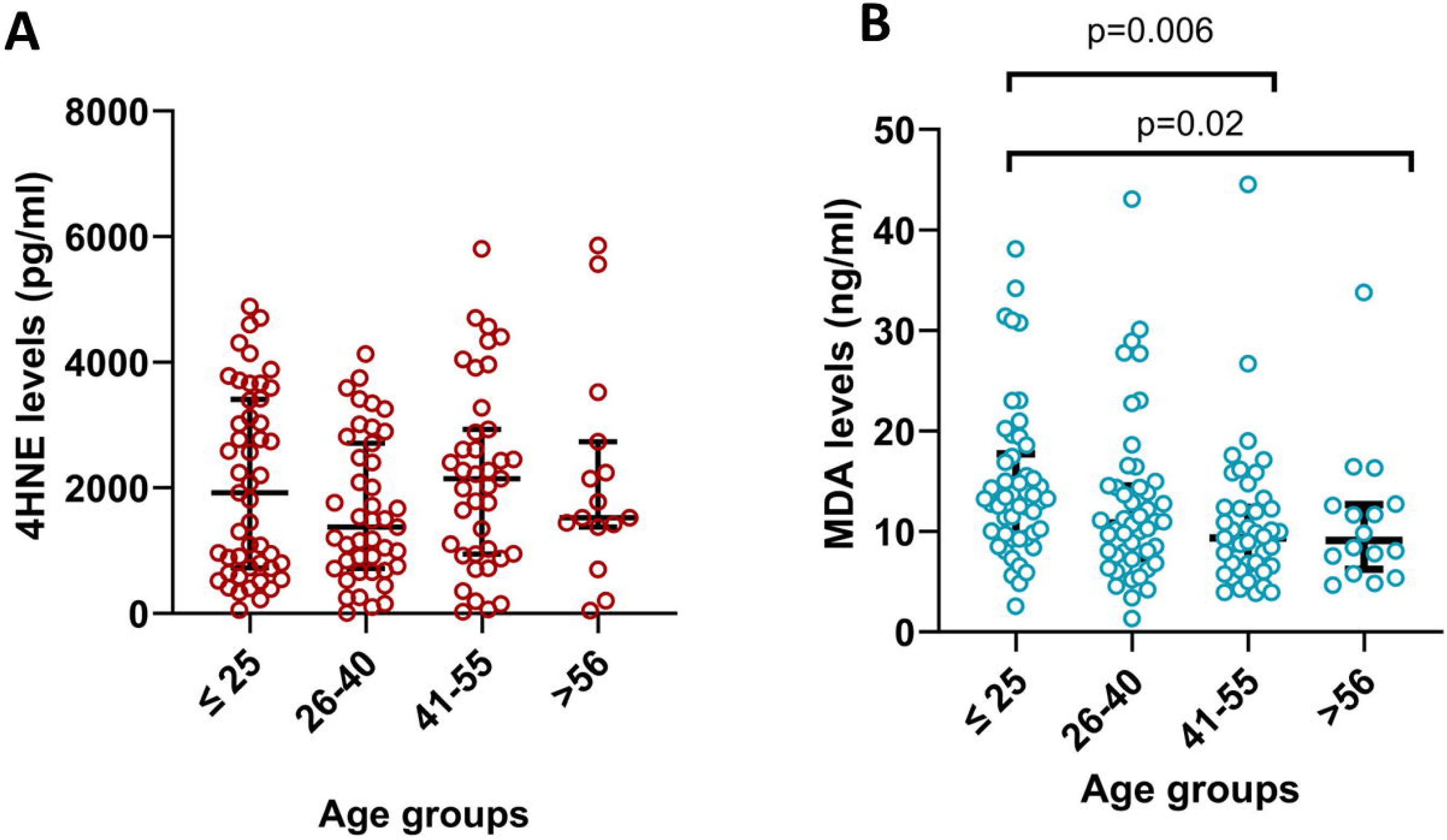
Relationship of markers of oxidative stress with age. We analyzed the levels of 4- HNE (dark red, A) and MDA levels (teal, B), in patients with acute dengue of different age groups. There were no differences in the 4-HNE levels in different age groups, whereas, MDA levels were significantly lower in those aged ≤ 25 years compared to those aged 41-55 years or those aged >56 years. The Mann-Whitney U test (two tailed) was used to calculate the differences between different cohorts. The error bars indicate the median and the interquartile ranges.

#### Association of ferritin levels with liver transaminase levels

As ferritin levels associate with clinical disease severity, we sought to understand the difference between ferritin levels in patients with varying liver transaminase levels. We found that the ferritin levels significantly increased with the rise in both AST and ALT levels (Fig 4A).

**Figure 4:**
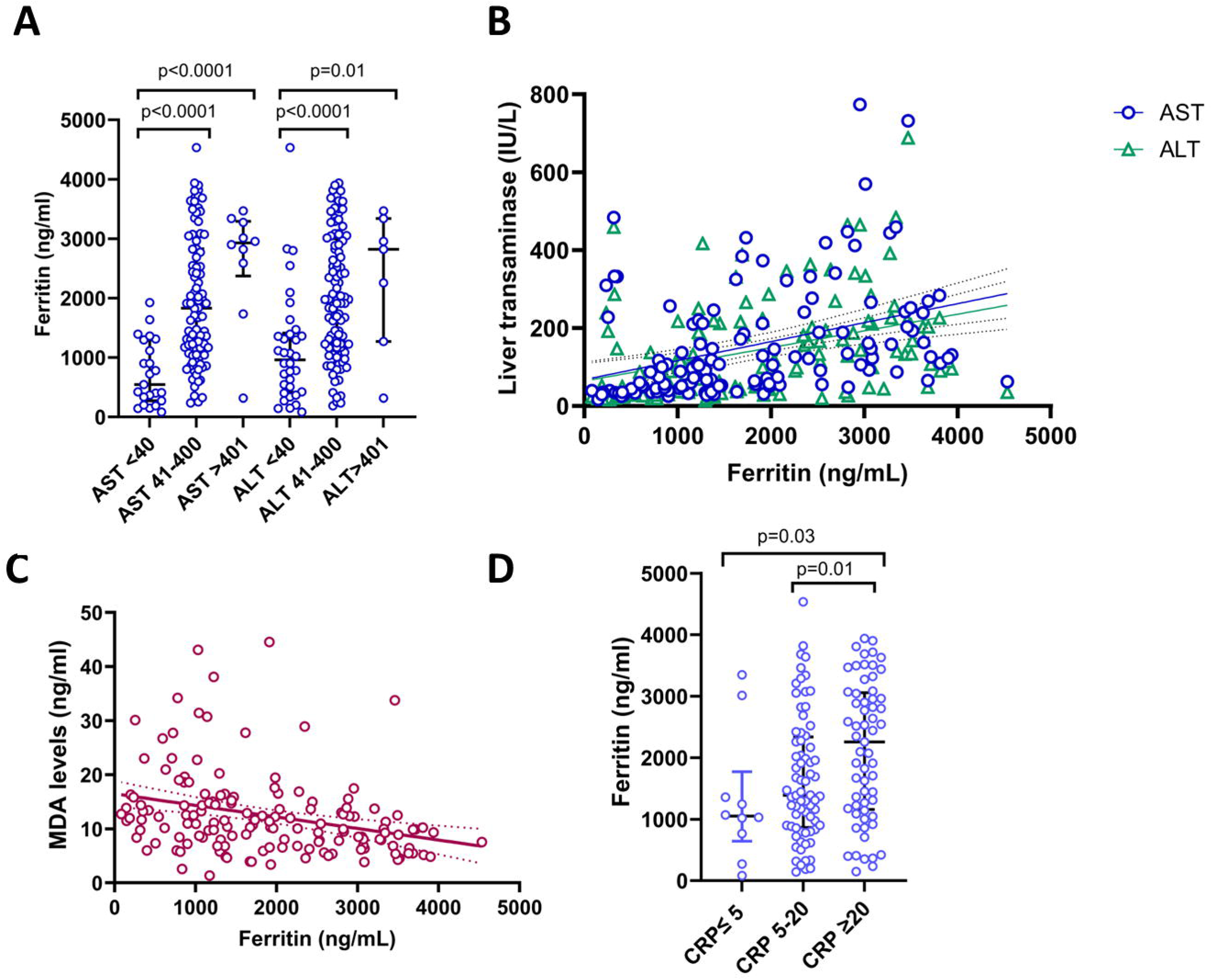
Association of ferritin levels with liver transaminase levels. Ferritin levels were compared in patients (n=151) with AST levels <40, 41-400 and >400 IU/L (A). Ferritin levels correlated with both AST and ALT (B), while ferritin levels inversely correlated with MDA levels (C). The ferritin levels were also assessed in patients with normal (≤5mg/ml), 5-20 and ≥20 mg/ml CRP levels. The Mann-Whitney U test (two tailed) was used to calculate the differences between different cohorts. The error bars indicate the median and the interquartile ranges. Spearman rank order correlation coefficient was used to evaluate the correlation between the liver transaminases, MDA and ferritin levels (two tailed).

Interestingly, all patients with a normal AST level had ferritin levels <2000 ng/ml. Ferritin levels correlated with both AST (Spearmans R=0.50, p<0.0001, Fig 4B), ALT levels (Spearmans R=0.55, p<0.0001, Fig 4B), CRP levels (Spearmans R=0.5, p<0.01), while inversely correlating with MDA levels (Spearmans R=-0.39, p<0.0001, Fig 4C). There was no association seen between ferritin and 4-HNE levels.

## Discussion

In this study we found that AST, ALT and CRP levels were significantly higher in patients with DHF compared to DF, as reported in many studies previously [6, 26]. Importantly, we found that AST, ALT, ferritin and CRP levels were significantly higher in those with central obesity and in overweight individuals. 45.3% of the patients were considered to have central obesity 45.4% were overweight, highlighting the increasing importance of obesity in many countries, including Sri Lanka. Although previous studies have shown that obesity was a risk factor for severe dengue, the association of obesity with an increase in liver transaminases, ferritin and CRP has not previously been described. Both ferritin and CRP associate with many other inflammatory mediators in dengue [34, 35], and ferritin levels >2000 ng/ml are though to reflect a ‘hyperinflammatory’ state [27]. Obesity has shown to be a risk factor for many viral infections [36], which has been attributed to dysfunctional adipose tissue, altered monocyte phenotypes, increase in mast cells with an altered phenotype, altered adipokine profile, increased intestinal permeability and endotoxaemia, chronic low-grade inflammation, dysfunctional NK and T cell responses and waning of neutralizing antibody responses [37–39], which could be relevant in the pathogenesis of severe dengue.

4-HNE and MDA are products of lipid peroxidation of omega-6 fatty acids, that are generated during oxidative stress [30]. 4-HNE is the main marker of lipid peroxidation and regulates many transcription factors that drive inflammation (NFкB), cell survival and proliferation (AP-1), energy regulation, including lipid and glucose homeostasis (PPAR) and those that maintain cellular redox homeostasis (Nrf2) [30]. Although MDA is also produced by lipid peroxidation and is elevated in many in those with different types of cancers, diabetes and neurodegenerative diseases, its function and generation is less studied than 4-HNE [30]. We found that 4-HNE significantly increased in those with high AST levels, and with ALT levels, although not significant. In contrast, MDA levels appeared to decrease with the rise in liver transaminases. Furthermore, while there was no difference in 4-HNE levels in obese and overweight individuals compared to lean individuals, MDA levels were significantly higher in lean individuals and in those with lower BMIs. Although 4-HNE levels have not been evaluated in patients with dengue previously, MDA levels assessed in a small cohort of patients, were found to be slightly lower in patients with severe dengue compared to those with non-severe dengue, on day 3 of illness [31]. Another study showed that there was no difference in MDA levels during early illness in patients with varying severity of illness [40]. However, as we found that MDA levels were significantly different in different age groups and in those with obese and overweight individuals, the ages and the presence of comorbidities are likely to affect the results. Furthermore, as Soundravally et al showed that the levels of MDA increased in the critical phase in patients with DHF, the timing of sample collection is also likely to affect the values. However, higher MDA levels were seen in early illness in younger and leaner patients, and since the MDA levels inversely correlated with the rise in liver transaminase levels and ferritin levels, higher MDA levels during early illness appear to associate with less severe disease, which should be further investigated.

As shown in previous studies, we too found that serum ferritin levels were significantly higher in patients with DHF compared to DF and also correlated with the levels of AST and ALT [27, 34]. Although ferritin was elevated in all patients with dengue, none of the patients with normal AST levels had ferritin levels >2000ng/ml, although some patients with normal ALT levels did have ferritin levels above this value. However, some patients with ferritin >2000ng/ml had normal CRP levels and had uncomplicated DF. Similarly, a significant number of patients who had AST levels and ALT levels <400 IU/L had ferritin levels >2000ng/ml. Therefore, although patients with ferritin levels >2000 ng/ml, are considered to have hyperinflammatory syndrome and severe dengue [27], this may not always be the case. Different studies have suggested lower and higher ferritin cut-off values as predictors of severe dengue, with varying sensitivity and specificity rates, which may be attributed to differences in clinical disease classification and methods of measuring ferritin levels [28, 41]. However, as some patients with mild uncomplicated dengue, with normal CRP levels did have ferritin levels of >2000ng/ml, it might be important to consider other parameters as well, in classifying patients as having hyperinflammatory syndrome.

## Conclusions

We found that obese and overweight patients were more likely to have elevated liver enzymes, and higher CRP and ferritin levels, suggesting that obesity and related metabolic disease is likely to contribute clinical disease severity and liver injury by inducing inflammation. 4-HNE, which is a marker of lipid peroxidation was found to be significantly elevated in patients with higher liver transaminase levels, while MDA was found to be higher in patients with milder disease and in younger and leaner individuals. Therefore, the different roles 4-HNE and MDA play in inducing liver injury should be further investigated. However, as 4-HNE does appear to associate with the extent of liver injury, it may be relevant to consider clinical trials with drugs such as N-acetyl cysteine for its effect on reducing oxidative stress and thereby liver damage, in addition to its reported antiviral effects [42].

## Data Availability

All data produced in the present work are contained in the manuscript.

## Acknowledgements

We are grateful to the NIH, USA (grant number 5U01AI151788-02) and the UK Medical Research Council.

